# Adult life-course trajectories of psychological distress and economic outcomes in midlife during the COVID-19 pandemic

**DOI:** 10.1101/2021.12.13.21267727

**Authors:** V Moulton, A Sullivan, A Goodman, S Parsons, G Ploubidis

## Abstract

This study used two British birth cohorts to examine whether pre-pandemic trajectories of psychological distress were associated with a greater risk of changes in financial and employment situation during the pandemic, as well as increased need for government support and use of other methods to mitigate their economic situation. We identified 5 differential life-course trajectories of psychological distress from adolescence to midlife and explored their relation to changes in financial and employment circumstances at different stages during the pandemic from May 2020 to March 2021, applying multinomial logistic regression and controlling for numerous early life covariates. In addition, we ran modified Poisson models with robust standard errors to identify whether different trajectories were more likely to have been supported by the benefit system, payment holidays, borrowing and other methods of mitigating the economic shock. We found that despite the UK governments economic response package economic inequalities for pre-pandemic psychological distress trajectories with differential onset, severity and chronicity across the life-course were exacerbated by the COVID-19 economic shock. Furthermore, the subsequent cut in government support, alongside increases in the cost of living may widen economic inequalities for differential pre-pandemic psychological distress trajectories, which in turn may also worsen mental health. This work highlights, different pre-pandemic trajectories of psychological distress were more vulnerable to economic shock.

## Introduction

At the start of the COVID-19 pandemic, there were warnings that those with prior mental health problems may face a greater risk of unemployment and financial hardship.^1,2^ Previous research on pandemics and emergencies (e.g., natural disasters) has shown that such events often widen health inequalities in society, and have a greater impact on socially disadvantaged groups.^3^ Additionally, financial adversity in times of economic recession has been shown to have disparate effects on individuals according to their prior mental health status,^4,5^ including an unemployment penalty associated with pre-existing psychological distress.^6^

The COVID-19 economic downturn differs from most recent recessions in that the effect on the labour market was immediate, the reduction in economic activity resulting from lockdown drove employers to reduce working hours or to temporarily or permanently terminate employees work in ‘non-essential’ occupations. In response, to insure households against economic shock in the short term and to mitigate sudden loss of income, the UK government introduced a number of economic measures. These included the Job Retention Scheme (JRS), whereby 80% of a furloughed employee’s wages (up to £2,500 per month) was paid by the government, and the Self-Employment Income Support Scheme (SEISS) grants explicitly contingent on having lost earnings. In addition, some social security payments, a £20 weekly increase to Universal Credit (UC) and the Working Tax Credit (WTC) were introduced, as well as the ability to apply for temporary payment deferrals on mortgages, rent, council tax, credit cards and personal loans.

Despite the UK governments response package, studies have found individuals were differentially exposed to the economic impact of the COVID shock in the UK, owing to individual characteristics, lower incomes, types of work and different private and public support mechanisms utilised.^7,8,9^ These factors could possibly have a greater impact on individuals with poorer pre-pandemic mental health. Emerging research from the COVID-19 pandemic suggests that mental health inequalities are being exacerbated. An investigation of 12 longitudinal studies in the UK general population, found that poor pre-pandemic mental health was associated with 5-13% greater odds of economic disruption, including loss of employment (OR=1.13 [1.06 to 1.21]) and income (OR=1.12 [1.06 to 1.19]) during COVID-19.^10^

Few studies have examined the influence of life-course mental health on economic outcomes, perhaps because of the bidirectional and mutually reinforcing nature of the relationship, as well as the availability of suitable data.^11,12^ However, a small body of work has focused on the long-term impact of child and adolescent mental health, on family income,^13,14^ unemployment,^15^ and earnings in adulthood. ^13,16^ In particular, Goodman et al. (2011) using data from the 1958 National Child Development Study (NCDS) find that psychological problems experienced by the age of 16 were associated with a 28% lower household income by age 50, while in the US Smith and Smith (2010) find that psychological problems in childhood are associated with a 35% reduction in adult family income.

Also, poor mental health during adulthood may shape future economic consequences, including vulnerabilities exacerbated by rapidly worsening economic conditions, such as those experienced during COVID-19. The age of onset of generalized anxiety disorder (median 24-50) and mood disorders (median 29-40)^17^ are often during the early stage’s of an adult’s work career and into the prime of their economic lives, and could therefore possibly hinder human capital accumulation.^18^ In addition, stigma and discrimination associated with mental health^19^ and an individual’s beliefs about their own abilities and decision making^20,21^ may further exacerbate adverse economic outcomes.

To the best of our knowledge, no study has investigated the relationship between differential mental health across the adult life-course and economic outcomes after the COVID-19 economic shock. In this study we examine trajectories of psychological distress from adolescence to midlife, thus giving an overview of the timing of onset along with the severity and chronicity at different life-stages. We investigate how these trajectories were associated with economic outcomes and changes in financial situations during the pandemic, thus distinguishing the relationship between the timing of onset and severity of pre-pandemic life-course psychological distress and adverse economic outcomes in later life as a result of the COVID-19 economic shock. We employ two large nationally representative British birth cohorts, the NCDS (1958) and BCS70 to examine whether,

1. after the economic shock distinct psychological distress trajectories were more at risk of changes in their financial and employment situation at three-time points during the pandemic,
2. and if they were more likely to have been supported by the governments COVID-19 economic mechanisms, as well as taking other measures to mitigate the change in their economic situation during the COVID-19 outbreak (March 2020 to March 2021).

## Methods

### Participants

Our data are from two ongoing cohort studies:

#### 1958 National Child Development Study

The NCDS follows the lives of 17,415 people that were born in England, Scotland or Wales in a single week in March 1958. The NCDS started in 1958 as the Perinatal Mortality Survey and captured 98% of the total births in Great Britain in the target week. The cohort has been followed up 10 times between ages 7 and 55.^22^

#### 1970 British Cohort Study

The BCS70 follows the lives of 17,198 people (representing 95% to 98% of the target population) born in England, Scotland and Wales people in a single week in April 1970. Participants have since been followed up nine times between ages 5 and 46.^23^

In addition, during the COVID-19 pandemic participants of the NCDS and BCS70 completed a web survey at three different time-points when they were aged 62 and 50, respectively. The first survey was conducted during the first national lockdown, between the 4 and 26 May 2020 (Wave 1: NCDS N: 5,178; BCS N: 4,223), the second survey was completed between the 10 September and 16 October (Wave 2: NCDS N: 6,282; BCS N: 5,320; when the first national lockdown had been lifted, but restrictions on social contact still remained, and the third survey was conducted during the third national lockdown, between the 1 February and 21 March 2021 (Wave 3: NCDS N: 6,757; BCS70 N:5,684).^24^

Our analytic sample included all participants in the NCDS and BCS70 surveys, excluding those who had died or emigrated by age 50 in the NCDS (n=15,291) and age 46 in the BCS70 n=17,486 (sample descriptives in Table S1 in the supplement). To deal with attrition and item non-response and to restore sample representativeness we used Multiple Imputation (MI) with chained equations, imputing 25 data sets^25^ separately for both cohorts at each wave of the COVID-19 surveys. All variables used in our main analysis, as well as a set of auxiliary variables (not in the substantial model of interest) were included in the imputation models to maximise the plausibility of the ‘missing at random’ (MAR) assumption in order to reduce bias due to missing data (Mostafa et al., 2021; Silverwood et al, 2020).^26,27^ As an additional sensitivity analysis all models were rerun in line with the ‘impute and delete’ method^28^ and the main findings did not differ (available on request).

### Measures

#### Trajectories of pre-pandemic psychological distress

Psychological distress was measured in both cohorts with the nine item version of the Malaise Inventory^29,30^ from ages 23,33,42 and 50 in the NCDS and ages 26,,34, 42 and 46 in the BCS70. Psychological distress captures depression and anxiety symptoms.^31^ In both surveys the Malaise items were assessed via written self-completion, either on paper or via computer. The Malaise inventory has been shown to have good psychometric properties,^32^ measurement invariance,^33^ and has been used in general population studies as well as investigations of high risk groups.^34^ In both cohorts, at age 16 four items from the Childrens’ Behavior Questionnaire reflective of affective disorders (Low mood, irritability, worry, and fearfulness), as reported by the child’s mother were employed.^30^ In prior work^35^ we used latent variable mixture models to identify five longitudinal typologies of psychological distress in both cohorts as the most parsimonious models.^36^

Figure 1a and 1b shows the means for each longitudinal latent class on each of the five measures (for results see Table S2a-c and Moulton et al., 2021). Although, the five trajectories were not identical in the two cohorts, some of the groupings were very similar. In both cohorts, the largest trajectory had few or no symptoms, ‘no symptoms’. Both cohorts also had a trajectory with persistent or repeated severe symptoms ‘stable-high symptoms’, and a trajectory with few symptoms in adolescence/early adulthood with ‘adult-onset’ (early 30’s) and favourable outcomes ‘adult-onset decreasing’. Both cohorts also had a trajectory with symptoms developing in midlife; in the NCDS the outcome was more positive ‘Midlife onset decreasing’ and in the BCS70 the symptoms remained severe ‘midlife-onset increasing’. The final trajectory in the NCDS was repeated minor symptoms ‘stable-low’ symptoms, while in the BCS70 the final trajectory had symptoms in early adulthood, but not in adolescence or midlife ‘early adult-onset decreasing’.

**Figure 1a:**
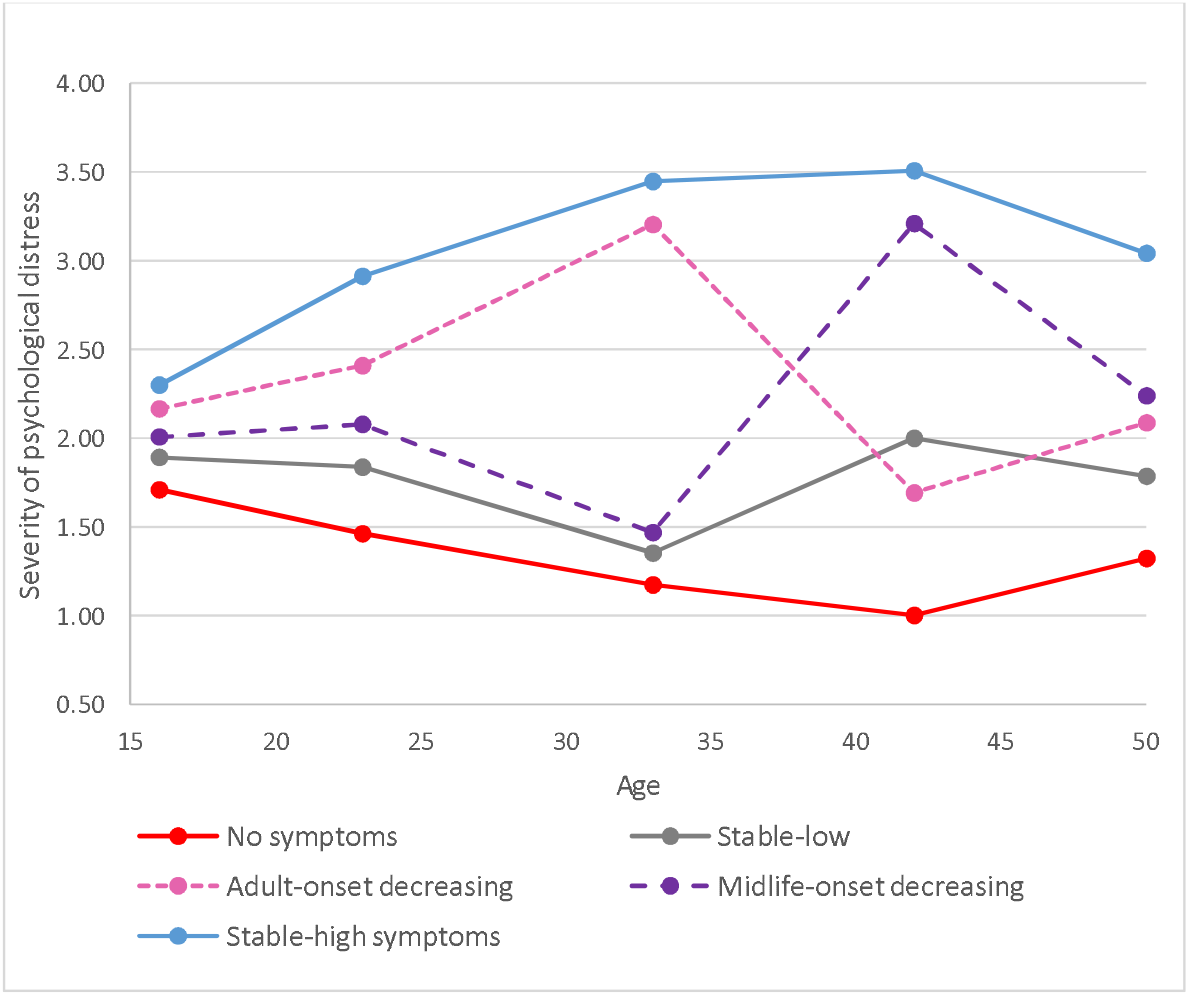
Five longitudinal classes of psychological distress from age 16 to 50 in the NCDS.

**Figure 1b:**
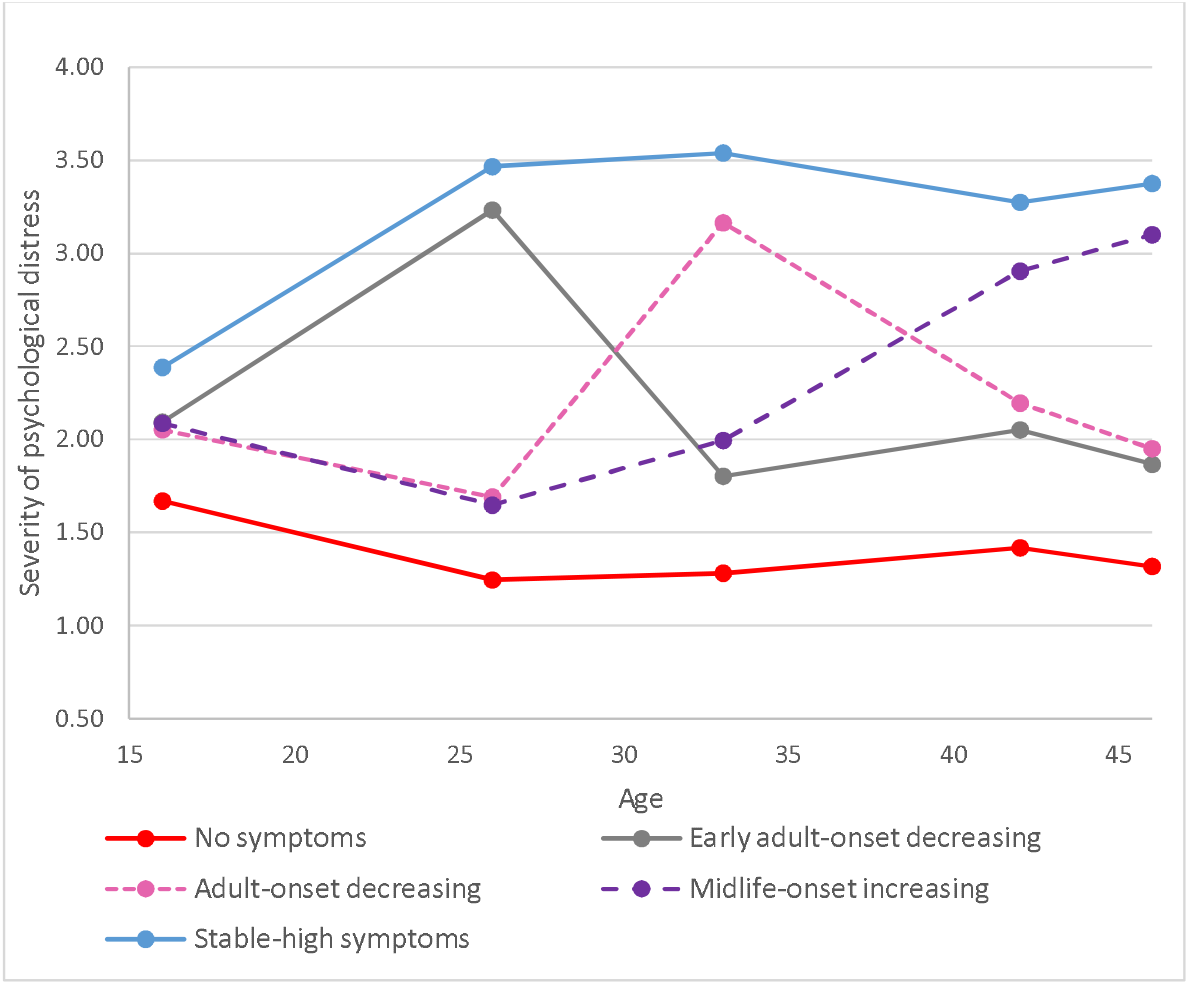
Five longitudinal classes of psychological distress from age 16 to 46 in the BCS70.

#### Outcomes

There were two outcomes capturing change in economic circumstances asked at three-time points during the COVID-19 pandemic; change in financial situation and change in employment circumstance from pre to during COVID-19. *Change in financial situation* was assessed, ‘overall how do you feel your current financial situation compares to before the Coronavirus outbreak’; much worse off, a little worse off, about the same (reference category) or a little/ much better off, recorded at three-time points during the pandemic. *Change in employment status* was derived by asking for economic status just before the first Coronavirus outbreak, as well as at three-time points during the pandemic. At each time point a variable was constructed to identify stability and change in employment throughout the pandemic (March to May 2020, March to Sept/Oct 2020, and March 2020 to Feb/March 2021) as follows; in work pre pandemic and stayed in work ‘work-work’, in work pre pandemic and was furloughed ‘work-furlough’, in work pre pandemic and was made unemployed ‘work-not work’ and other groups (e.g. no change ‘retired-retired’, ‘unemployed-unemployed’ and change, e.g. ‘work-retired’).

In addition, we captured a variety of methods used to mitigate the economic shock: made any new benefit claims, taken payments holidays since the outbreak, borrowed, reduced consumption or used savings. *New benefit claims* were assessed at each wave by asking, ‘since the Coronavirus outbreak have they or their partner made any new claims for any of the following: free school meals, universal credit, Employment and Support Allowance (ESA), Statutory Sick Pay (SSP), council tax support or reduction, carers allowance or personal independent payments, new governmental financial support for self-employed people, or not. These were combined to create a single variable, any new claims (1) from March 27^th^ 2020 to 21 March 2021, or not (0). *Payment holidays* was examined by asking at each wave ‘since the coronavirus outbreak had they used any mortgage, rent, council tax, or interest payment holidays or other debt repayments’. These were amalgamated into a single variable, taken any payment holidays from the end of March 2020 to March 2021 (1), or not (0). Also, *increase in financial help* was assessed by asking at wave 2 and 3 ‘since the Coronavirus outbreak in March 2020, have (they or their partner) received financial help, in the form of money or by paying for goods (for example groceries, medicines) from…family and friends’, and if so was this an increase since the outbreak in March 2020. These questions were combined into a single variable, financial help increased from the end of March 2020 to March 2021 (1), or not (0). Other mitigation strategies were examined by asking ‘You said that you are worse off now compared to before the Coronavirus outbreak in March 2020. Have you (or your partner) done any of the following as a result of this…reduced spending, used savings, new borrowing from bank or credit card, new borrowing from family and friends’. These were transformed into dichotomous variables; reduced spending (1), or not (0); used savings (1) or not (0); new borrowing from bank or credit card (1) or not (0), and new borrowing from family and friends (1) or not (0) from the end of March 2020 to March 2021. Also, in order to examine the potential impact of the removal of the £20 UC uplift we created a variable ‘claiming universal credit or not’ based on all making a UC claim from January 2020 to March 2021.

#### Potential confounders

We include in our analysis a rich set of variables comprising early life factors (sex, ever breastfed, mother smoked daily during pregnancy, gestation period, and birthweight), socio-economic factors (parental social class, education, housing tenure, access to amenities, total household income, crowding and marital status), parental factors (maternal age at birth, mother worked at all in first five years, separated from child and read to), child behavior and health (cohort member bedwetting since age 5, had any medical conditions, Body Mass Index (BMI)), and cognitive ability (details are available in supplementary Table S3).

#### Analytic approach

To answer question 1, we used the psychological distress profiles to explore their relation to changes in financial and employment circumstances at three timepoints during the Coronavirus outbreak, applying multinomial logistic regression and controlling for numerous pre-adult covariates. In addition, we ran modified Poisson models with robust standard errors that return risk ratios for ease of interpretation and to avoid bias due to non-collapsibility of the odds ratio^37^ to identify whether differential life-course trajectories of psychological distress were more likely to have been supported by the benefit system, payment holidays, borrowing and other methods of mitigating the economic shock during the pandemic, thereby answering question 2.

## Results

### Pre-pandemic psychological distress trajectories

In our analytic sample, in the NCDS 40.9% had ‘no symptoms’ of psychological distress across the life-course, while 19.9% had ‘stable-high’ symptoms, 16.4% ‘stable-low’, 11.9% ‘midlife-onset’ decreasing, and 10.8% ‘adult-onset’ decreasing. In the BCS70, half (52.4%) had ‘no symptoms’, 20.4% ‘stable-high symptoms’, 10.9% ‘adult-onset’ decreasing, 8.8% ‘early-adult onset’ decreasing, and 7.6% ‘midlife-onset’ increasing.

### Descriptives of psychological distress trajectories and financial circumstances during the COVID-19 pandemic

As shown in Table 1 during the pandemic, a high proportion of participants’ financial situations were similar to pre-pandemic circumstances; over half (ranging from 59.5% to 51.4%) in the NCDS and fluctuating from 42.5% to 53.4% in the BCS70. However, any change in circumstances from pre to during the pandemic was greater for those with worsening finances, with a smaller number improving their financial circumstances during the crisis. For both cohorts, throughout the pandemic a higher proportion of the ‘no-symptoms’ trajectory were financially better off than pre-COVID, compared to the ‘stable-high’ symptoms trajectory. Other changes in financial circumstances were broadly the same across differential psychological distress trajectories, until Sept/Oct 2020 when a higher proportion of the ‘stable-high’ symptoms trajectory in the BCS70 were much worse off financially than pre-COVID, compared to the ‘no symptoms’ trajectory (18.4% compared to 8.1% respectively). This pattern was replicated in spring 2021, in addition a higher proportion of ‘midlife-onset’ in both cohorts as well as ‘stable-high’ in the NCDS were in a much worse financial situation than over a year ago pre-COVID 19.

**Table 1:**
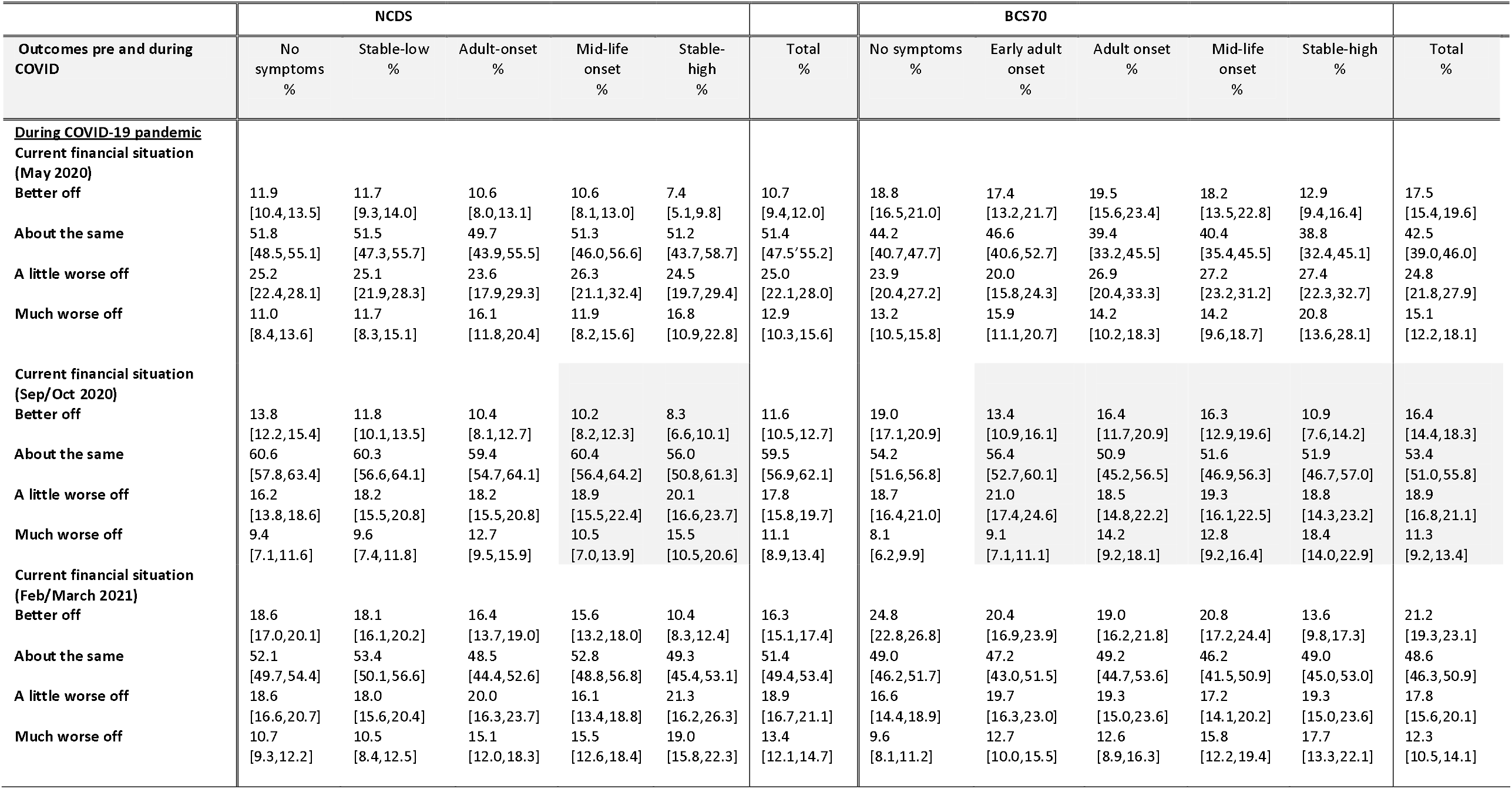
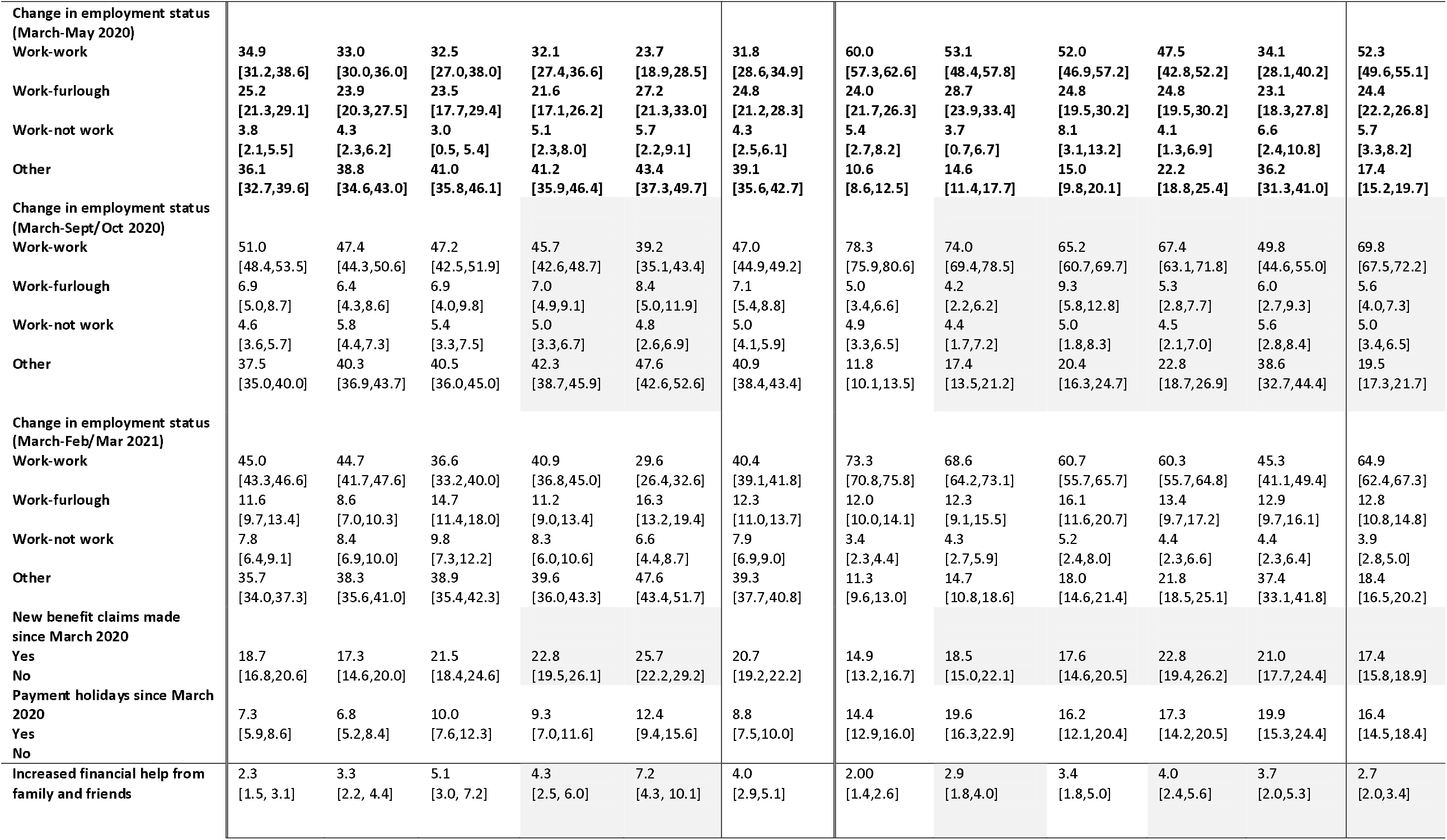
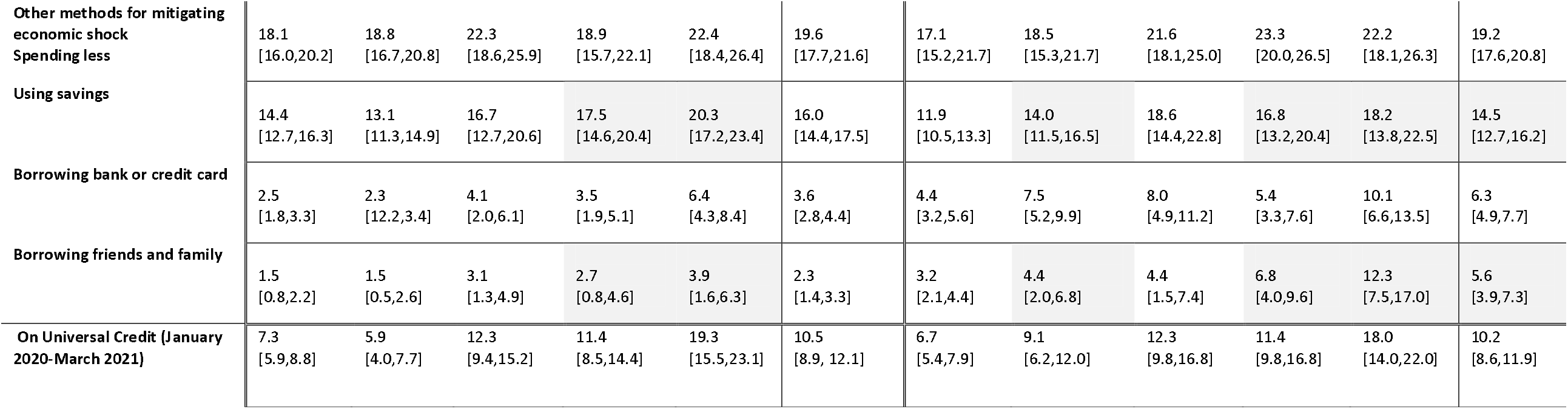
Distribution of cohort members financial circumstances during the COVID-19 crisis by pre-pandemic psychological distress trajectories.

Overall, at the beginning of the pandemic a quarter of participants in the NCDS and BCS70 were put on furlough, by early Autumn this had reduced to 7.1% and 5.7% for the NCDS and BCS70 respectively, and increased slightly in the Spring of 2021, while a few, an eighth had lost their jobs. For those working, there was no difference in the proportion who were furloughed or lost their jobs during the pandemic by distinct psychological distress trajectories.

Overall, 20.7% of the NCDS and 17.4% of the BCS70 made new benefit claims, and 16.4% of the BCS70 and 8.8% of the NCDS took payment holidays. However, a higher proportion of the ‘stable-high’ trajectory in the NCDS claimed for new benefits and took payment holidays, compared to the ‘no-symptoms’ trajectory. Likewise, in the BCS70 a higher proportion of the ‘stable-high’ symptoms trajectory, along with the ‘midlife-onset’ trajectory, than the ‘no-symptoms’ trajectory made new benefit claims. Throughout the pandemic, a higher proportion of the ‘adult-onset’, midlife-onset and ‘stable-high’ trajectories, compared to the ‘no symptom’ trajectory in both cohorts were claiming Universal Credit. Overall, a higher proportion of cohort members, reduced consumption or used savings during the pandemic to mitigate the financial shock, while a smaller proportion borrowed from family or formal institutions. However, compared to the ‘no-symptoms’ trajectory, a higher proportion of the ‘stable-high’ trajectories in both cohorts borrowed from banks or credit cards, and received financial help or borrowed from family and friends.

### Risk of change in economic outcomes during the course of the pandemic

Table 2 presents the relative risks in the fully adjusted models, associated with each of the economic outcomes during the pandemic for different trajectories of psychological distress, with the largest trajectory ‘no symptoms’ used as the reference category in the analysis.

**Table 2:**
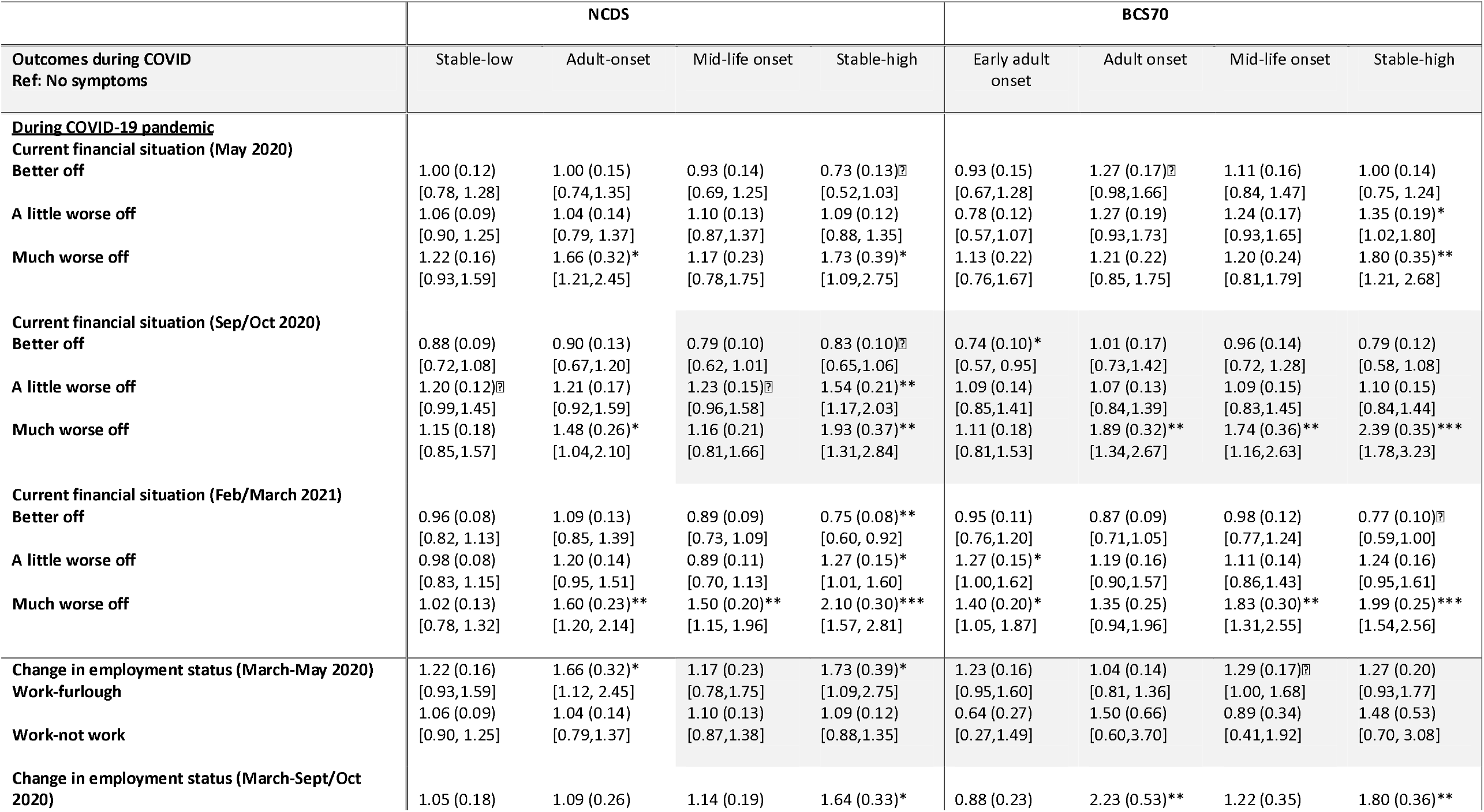
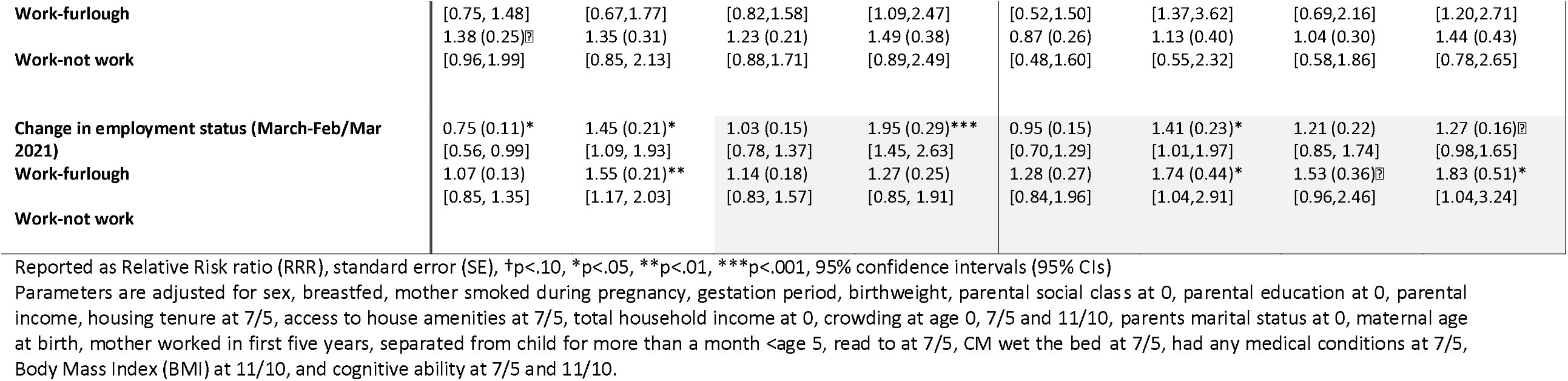
Relative risk (RR) of change in financial and employment circumstances associated with pre-pandemic psychological distress trajectories in the NCDS and BCS70 during the COVID-19 pandemic.

#### Change in financial circumstances during the course of the pandemic

Throughout the pandemic, the ‘stable-high’ trajectory in both the NCDS [W1: RRR=1.7, 95% CI 1.1-2.8; W2: RRR= 1.9, 95% CI 1.3-2.8; W3: RRR=2.1, 95% CI 1.6-2.8] and the BCS70 [W1: RRR=1.8, 95% CI 1.2-2.7; W2: RRR=2.4, 95% CI 1.8-3.2; W3: RRR=2.0, 95% CI 1.5-2.6], were at greater risk of being much worse off financially. In contrast, a year into the pandemic the ‘no-symptom’ trajectory, than the ‘stable-high’ trajectory [RRR: 0.75, 95% CI: 0.6-0.9] in the NCDS were associated with being better off financially at the beginning of 2021. Also, in the NCDS ‘adult-onset’ [W1: RRR=1.7, 95% CI 1.2-2.5; W2: RRR=1.5, 95% CI 1.2-2.5; W3: RRR=1.6, 95% CI:1.2-2.1] was associated with a worsening financial situation throughout the pandemic. Other pre-pandemic psychological distress trajectories were also associated with a greater risk of being financially worse off at different stages during the pandemic; in the early autumn 2020 the BCS70 ‘adult-onset’ (RRR= 1.9, 95% CI 1.3-2.7) and ‘midlife-onset’ (RRR= 1.7, 95% CI 1.2-2.6), and by Spring 2021 in both cohorts the ‘midlife-onset’ (BCS70: RRR=1.8, 95% CI 1.3-2.6; NCDS RRR= 1.5, 1.2-2.0), and in the BCS70 ‘early-adult onset’ (RRR= 1.4, 95% CI 1.1-1.9). The worsening of financial circumstances related to between 1 in 7 to 1 in 5 of the trajectories with prior psychological distress, compared to around a tenth of the ‘no-symptoms’ trajectory (details are available in supplementary Table S5).

#### Change in employment during the course of the pandemic

During the pandemic, if in work prior to the COVID-19 outbreak both the ‘adult-onset’ and ‘stable-high’ trajectories were related to a greater risk of furlough. And by Spring 2021 the ‘adult-onset’ trajectory in both cohorts [NCDS RRR=1.6, 95% CI 1.2-2.0; BCS70 RRR=1.7, 1.0-2.9], and the ‘stable-high’ in the BCS70 [RRR=1.8, 95% CI 1.0-3.2], were also associated with a greater risk of unemployment.

### Government support and other approaches to mitigate the economic shock

In both cohorts (Table 3), the ‘stable-high’ trajectory was associated with around a 30% increase in the risk of making new benefit claims, likewise the ‘midlife-onset’ trajectory were more likely to access government support (NCDS: RR=1.2, 95% CI 1.0-1.4; BCS70 RR=1.5, 95% CI 1.3-1.8). Also, the ‘stable-high’ trajectory had a 72% in the NCDS and 30% in the BCS70 risk of taking payment holidays during the pandemic (representing around 12.4% in the NCDS and 18.3% in the BCS70 of the stable-high trajectory – see supplementary Table S5).

**Table 3:** Relative risk (RR) of using methods to mitigate the economic shock associated with pre-pandemic psychological distress trajectories in the NCDS and BCS70 during the COVID-19 pandemic.

|  | NCDS |  |  |  | BCS70 |  |  |  |
| --- | --- | --- | --- | --- | --- | --- | --- | --- |
| Outcomes during COVID<br>Ref: No symptoms | Stable-low | Adult-onset | Mid-life onset | Stable-high | Early adult onset | Adult onset | Mid-life onset | Stable-high |
| **Any new benefit claims March 2020 – March 2021** | 0.95 (0.08) | 1.11 (0.11) | 1.21 (0.10)* | 1.31 (0.11)** | 1.18 (0.12) | 1.15 (0.11) | 1.48 (0.13)*** | 1.34 (0.11)** |
| Yes | [0.81, 1.12] | [0.92, 1.35] | [1.02, 1.42] | [1.11, 1.55] | [0.96, 1.45] | [0.96, 1.39] | [1.25, 1.76] | [1.13, 1.58] |
| No |  |  |  |  |  |  |  |  |
| **Payment holidays since March 2020 – March 2021** | 0.98 (0.12) | 1.37 (0.19)* | 1.29 (0.17)▯ | 1.72 (0.23)*** | 1.32 (0.11) | 1.09 (0.14) | 1.19 (0.12)▯ | 1.30 (0.13)* |
| Yes | [0.77, 1.26] | [1.03, 1.82] | [0.99, 1.69] | [1.31, 2.25] | [1.12, 1.57] | [0.84, 1.40] | [0.98, 1.44] | [1.06, 1.59] |
| No |  |  |  |  |  |  |  |  |
| **Increased financial help from family and friends** | 1.45 (0.29)▯ | 2.05 (0.42)** | 1.80 (0.40)* | 2.85 (0.60)*** | 1.36 (0.27) | 1.69 (0.37)* | 1.86 (0.40)** | 1.72 (0.42)* |
|  | [0.98, 2.15] | [1.36, 3.10] | [1.06, 2.80] | [1.87, 4.35] | [0.92, 2.00] | [1.09, 2.61] | [1.22, 2.84] | [1.05, 2.81] |
| **Other methods for mitigating economic shock** | 1.07 (0.08) | 1.27 (0.11)** | 1.06 (0.09) | 1.31 (0.10)** | 1.09 (0.11) | 1.30 (0.15)** | 1.38 (0.12)*** | 1.36 (0.19)*** |
| Spending less | [0.93, 1.24] | [1.06, 1.53] | [0.97, 1.24] | [1.12, 1.53] | [0.89, 1.35] | [1.09, 1.54] | [1.16, 1.64] | [1.08, 1.72] |
| **Using savings** | 0.93 (0.08) | 1.17 (0.13) | 1.23 (0.11)* | 1.46 (0.11)*** | 1.23 (0.14)▯ | 1.60 (0.16)*** | 1.46 (0.18)** | 1.59 (0.19)*** |
|  | [0.78, 1.12] | [0.94, 1.45] | [1.01, 1.51] | [1.25, 1.70] | [0.98, 1.54] | [1.30, 1.97] | [1.14, 1.86] | [1.25, 2.02] |
| **Borrowing bank or credit card** | 0.97 (0.24) | 1.62 (0.44)▯ | 1.41 (0.41) | 2.59 (0.51)*** | 1.68 (0.32)** | 1.71 (0.30)** | 1.19 (0.26) | 1.93 (0.39)** |
|  | [0.59, 1.59] | [0.93, 2.82] | [0.78, 2.55] | [1.74, 3.83] | [1.15, 2.47] | [1.20, 2.44] | [0.76, 1.86] | [1.28, 2.90] |
| **Borrowing friends and family** | 0.97 (0.38) | 1.98 (0.70)▯ | 1.62 (0.55) | 2.44 (0.70)** | 1.27 (0.36) | 1.18 (0.39) | 1.94 (0.39)** | 2.70 (0.59)*** |
|  | [0.44, 2.15] | [0.97, 4.03] | [0.82, 3.22] | [1.37, 4.35] | [0.71, 2.27] | [0.61, 2.30] | [1.30, 2.92] | [1.73, 4.22] |
| **On universal credit (Jan 2020-March2021)** | 0.83 (0.13) | 1.47 (0.16)** | 1.45 (0.19)** | 2.10 (0.25)*** | 1.18 (0.19) | 1.61 (0.20)*** | 1.47 (0.21)** | 1.64 (0.16)*** |
|  | [0.61, 1.23] | [1.18, 1.84] | [1.12, 1.88] | [1.65, 2.66] | [0.86, 1.63] | [1.25, 2.07] | [1.11, 1.97] | [1.35, 1.98] |

Reported as Relative Risk (RR), standard error (SE), †p<.10, \*p<.05, \*\*p<.01, \*\*\*p<.001, 95% confidence intervals (95% CIs)

The ‘stable-high’ trajectory were also at a higher risk of using a variety of private methods to mitigate the economic shock, including reducing consumption, using savings, receiving financial help from family and borrowing. In particular, the NCDS ‘stable-high’ trajectory were associated with a 2.8 fold risk of relying on financial help from relatives, as well as 2.6 times the risk of borrowing from a bank or using credit cards. Similarly, the ‘stable-high’ trajectory in the BCS70 were associated with the risk of borrowing, from family (RR= 2.7, 95% CI 1.7-4.2) and financial institutions (RR= 1.9, 95% CI 1.3-2.9).

Other differential trajectories of psychological distress, compared to ‘no-symptoms’ were also more at risk of using other methods to supplement their worsening financial situation during the pandemic. In the NCDS the ‘adult-onset’ trajectory were associated with a 105% increase in receiving financial help from family, and a 37% increase in taking payment holidays, and a 27% increase in risk of a reduction in spending; while the ‘midlife-onset’ trajectory also relied on family financial support (RR=1.8, 95% CI 1.1-2.8), as well as using their savings (RR= 1.2,95% CI 1.0-1.5). In the BCS70, the ‘adult-onset’ and ‘midlife-onset’ trajectories were more at risk of both reducing consumption (RR= 1.3, 95% CI 1.1-1.5; RR=1.4, 95% CI 1.2-1.6) and spending their savings (RR=1.6, 95% CI 1.3-2.0; RR= 1.5, 95% CI 1.1-1.9). In addition, both trajectories were at increased risk of receiving financial support from family, while the ‘adult-onset’ trajectory were also associated with a 71% increase, as well as the ‘early-adult onset’ a 68% increase in risk of borrowing from banks and credit cards. Within each of the trajectories a higher proportion were making new benefit claims, reducing consumption, using savings and taking payment holidays to mitigate the economic shock, a lower proportion were borrowing or increasing financial support from friends (See Table S5).

From January 2020 to May 2021, as shown in Figure 2a and 2b the ‘adult-onset’, midlife-onset’ and ‘stable-high’ trajectories were more likely to be claiming Universal Credit and therefore more at risk of impact from the loss of the temporary £20 Universal Credit uplift.

**Figure 2:**
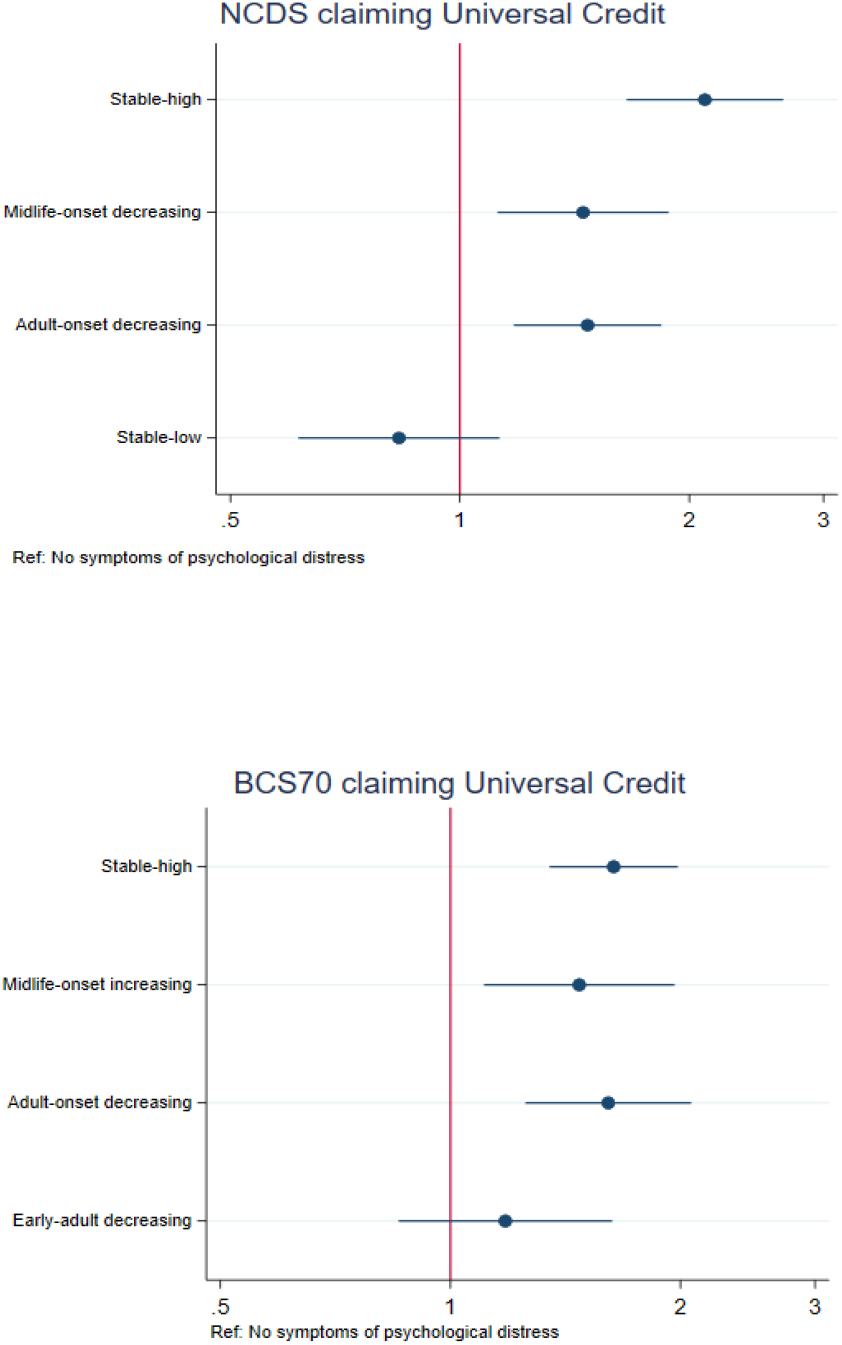
Relative risk of claiming Universal Credit January 2020 to Feb/March 2021 Parameters are adjusted for sex, breastfed, mother smoked during pregnancy, gestation period, birthweight, parental social class at 0, parental education at 0, parental income, housing tenure at 7/5, access to house amenities at 7/5, total household income at 0, crowding at age 0, 7/5 and 11/10, parents marital status at 0, maternal age at birth, mother worked in first five years, separated from child for more than a month <age 5, read to at 7/5, CM wet the bed at 7/5, had any medical conditions at 7/5, Body Mass Index (BMI) at 11/10, and cognitive ability at 7/5 and 11/10.

## Discussion

Pre-pandemic psychological distress was related to the risk of worsening financial circumstances during the pandemic. This finding held across a range of life-course psychological distress trajectories In both cohorts the ‘stable-high’ symptoms group were financially worse off at different stages during the pandemic. By, March 2021 most trajectories with prior symptoms of psychological distress across the life course irrespective of age of onset, severity, and proximal occurrence were associated with a greater relative risk of a worsening financial situation. For those in employment pre-pandemic, two trajectories in both cohorts, the ‘adult-onset’ (early 30’s) and the ‘stable-high’ were related to changes in their employment situations. Throughout the pandemic both trajectories were supported by the JRS scheme, however by March 2021 they were also associated with a greater relative risk of unemployment. The ‘stable-high’ symptoms trajectories were also supported by the benefits system, they along with the ‘midlife-onset’ trajectories were related to a greater relative risk of making a new claim during the pandemic. In addition, in both cohorts, from January 2020 to March 2021, the ‘adult-onset’, ‘midlife-onset’ and ‘stable-high’ trajectories were associated with claiming Universal Credit, and therefore receiving the £20 weekly uplift. Despite the government response, the worsening financial situations for pre-pandemic psychological distress trajectories were also associated with other methods of mitigation. While most psychological distress trajectories were related to a greater relative risk of using private means by reducing spending, dis-saving and relying on increased financial help from family and friends, the ‘stable-high’ symptoms trajectories in particular (along with a greater risk of borrowing from banks or credit cards for the ‘early-adult’ and ‘adult-onset’ trajectories in the BCS70) were associated with alleviating their financial circumstances through taking payment holidays and borrowing from institutions and friends.

At an aggregate level, the economic policy measures taken by the government, especially the JRS and UC uplift, were successful in broadly insuring households against the economic shock.^38^ Indeed, in this study during the pandemic differential psychological distress trajectories with symptoms pre-pandemic were increasingly likely to have been supported by the JRS and benefit system. However, despite this support most psychological distress trajectories with pre-pandemic psychological distress were associated with a relative risk of worsening financial circumstances. The Coronavirus SEISS and JRS ceased from the 30^th^ September 2021, and at the time of writing despite calls against stopping the £20 UC uplift, it also concluded on the 6^th^ October 2021. Removal of this support could be particularly detrimental for pre-pandemic psychological distress trajectories. In addition, this loss of financial support will be compounded by the increased cost of living including high energy costs, increased National Insurance Contributions in April 2022,^39^ and future economic uncertainty over COVID-19, Brexit, the national debt, and climate change, which may further exacerbate economic inequalities for these trajectories.

As with other studies, we find that individuals were differentially exposed to the economic impact of COVID in the UK,^7,8,9^ and specifically individuals with poor pre-pandemic mental health.^10^ Although, ‘adult-onset’ decreasing and ‘stable-high’ symptom trajectories were associated with being supported by the furlough scheme throughout the pandemic, by spring 2021 they were also related to a greater relative risk of unemployment. There are a number of mechanisms, (beyond the scope of this study to examine), which might explain why these two life-course psychological distress trajectories were associated with employment changes during the pandemic. For example, both trajectories were related to higher levels of psychological distress in their early thirties, which may coincide with an important period of employment transition,^40^ thus reducing human capital accumulation and skill acquisition^12^ which in turn influences future employment outcomes. During COVID-19, the labour market shock was heterogenous, industries which involved contact with people and ‘elementary’ occupations were hardest hit^8^ as well as low earners.^9^ Also, studies suggest there is evidence of discrimination against applicants with a history of mental health problems^41^ and a greater impact of job loss during recessions.^5^

In this study, psychological distress trajectories with prior symptoms were associated with greater relative use of different mitigation strategies, including dis-saving, and reduced consumption. Particularly worrying was the association between the ‘stable-high’ symptoms trajectory in both cohorts and in the BCS70 the ‘early-adult onset’ and ‘adult-onset’ trajectories with using methods such as payment holidays, and borrowing from friends and institutions, albeit on aggregate within trajectory the least likely approaches compared to using other mitigation methods such as making benefit claims. If borrowing and payment holidays are not short-term solutions to the economic crisis, they could lead to problem debt. A meta-analysis of pooled odds ratios showed a significant relationship between debt and mental disorder (OR = 3.24) and depression (OR=2.77).^42^ In prior studies indebtedness or an increase in debt levels was associated with subsequently poorer mental,^43^ and more severe the debt being related to more severe health difficulties.^42^ Previous work has also shown the relationship between low income and mental health is largely mediated by debt.^44,45^

The relationship between mental health and economic hardship is complex and contentious, whether the relationship is better explained by social causation, social selection or both.^12,46^ In this study we investigated whether the consequences of life-course psychological distress trajectories after an unprecedented economic shock were related to poorer financial and employment outcomes. Although this was an abrupt event, we cannot rule out the influence of prior financial circumstances, both as a confounder and as an aggravation of economic inequalities for trajectories with prior poor mental health. We conducted further analysis (Table S6-S8) to examine how trajectories with prior psychological distress were managing financially prior to the COVID-19 pandemic. Compared to the ‘no-symptoms’ trajectory none were associated with living comfortably, and the trajectories with higher proximal symptoms of psychological distress were at a greater relative risk of financial difficulties pre-pandemic. We stratified the samples into financially comfortable and financially struggling pre-pandemic and found that the risk of being worse off during COVID-19 for the ‘stable-high’ trajectories was associated with both those struggling, as well those who were financially comfortable pre-pandemic. In addition, psychological distress trajectories with onset at different stages in the life-course (‘adult-onset’ and ‘midlife-onset’) who were comfortable pre-pandemic were at risk of a worsening financial situation. This indicates both a possible increase in economic inequalities, as well as a likely vulnerability for some adult life-course psychological distress trajectories to economic shocks.

## Strengths and limitations

Our study is the first to relate long-term individual longitudinal psychological distress data before the COVID-19 pandemic to economic outcomes during the COVID-19 pandemic. Other unique strengths include nationally representative samples, large sample sizes, prospective follow-up from birth to midlife, and economic data collected at three discrete time points during the COVID-19 pandemic from May 2020 to March 2021.

There are a number of limitations of this study. Our findings can only be generalised to those born in Britain in 1958 and 1970 or close to those years. As with most longitudinal research, selective attrition has occurred. However, we included auxiliary variables in the multiple imputation models, including mental health and related variables from birth, which allows for predicting missing data with greater accuracy and minimizing non-random variation in these values.^47^ Also bias due to unmeasured confounding especially time varying confounding, in the observed association between the pre-pandemic psychological distress trajectories and economic outcomes during the pandemic cannot be ruled out. In terms of the method adopted, latent classes are approximations of symptom patterns in the data and do not represent actual data points, but are evidenced based summaries of psychological distress in the cohorts (see appendix S2 for more limitations on this approach). Also, relationships between subjective measures of poor financial situation and depression can arise irrespective of ‘objective’ measures of financial situation, therefore indicating a person-specific effect.^48^ However, in this study the measures of psychological distress and economic outcomes were time variant, and we investigated both subjective measures of financial circumstance, as well as ‘objective’ measures of employment situation and mitigation methods during COVID-19. This study is based on data collected until spring 2021, and therefore we do not know if the financial and employment situations for these psychological distress trajectories, stayed the same, worsened or improved prior to the removal of government support in early autumn 2021.

## Conclusions

Economic inequalities for pre-pandemic psychological distress trajectories with differential onset, severity and chronicity across the life-course seem to have been exacerbated by the COVID-19 economic shock. In addition, the ‘adult-onset’, ‘midlife-onset’ and the ‘stable-high’ symptoms trajectories who were living comfortably prior to COVID-19 were also disproportionately at risk of poorer economic outcomes. During the pandemic life-course psychological distress trajectories with prior symptoms were more likely supported by the governments’ economic response package, however the subsequent cut in support, alongside ‘post-pandemic’ increases in the cost of living especially high energy bills, and for example, the rise in employee National Insurance contributions in April 2022 will put further strain on the finances of those that have experienced psychological distress over their life course. The bi-directional nature of mental health inequality could in turn increase psychological distress symptoms or reduce the chances and delay improvements in current and future mental health for these trajectories. Highlighting the different mental health trajectories across the life-course which are vulnerable to economic shock and likely in need of further financial and mental health support is crucial. More research is needed to explore the possible mechanisms throughout the adult life-course related to vulnerability to labour market and economic shocks, as well as monitor these trajectories over the short and medium turn, especially in relation to employment opportunities, debt problems and increased mental health symptomatology.

## Data Availability

All data produced in the present study are available upon reasonable request to the authors

